# Gestational diabetes mellitus and hypertension causally increased the risk of postpartum depression: a Mendelian randomization analysis

**DOI:** 10.1101/2024.12.14.24319038

**Authors:** Jia Jia, Haojie Liu, Liying Cao, Ying Xing

## Abstract

**Background:** Observational studies have presented inconsistent findings on the association between gestational diabetes mellitus, gestational hypertension, and postpartum depression. This study used Mendelian randomization to examine the potential causal relationship between gestational diabetes mellitus, gestational hypertension, and postpartum depression.

**Methods:** We obtained data from genome-wide association study databases. Single nucleotide polymorphisms associated with gestational diabetes mellitus (5,687 cases; 117,892 controls), gestational hypertension (7,686 cases; 115,893 controls), and postpartum depression (7,604 cases; 59,601 controls) were analyzed. Various Mendelian randomization methods were applied, including inverse variance weighted, weighted median, MR-Egger, simple mode, and weighted mode. Sensitivity analyses such as the MR-Egger intercept test, funnel plot, MR-PRESSO analysis, Cochran’s Q test, and leave-one-out tests confirmed the robustness of our findings. The MR-Steiger test was applied to verify the causal direction from exposure to outcome.

**Results:** Genetically predicted gestational diabetes mellitus was significantly associated with increased postpartum depression risk (IVW OR = 1.09; 95% CI: 1.03- 1.14; p = 1.24×10⁻³), as was gestational hypertension (IVW OR = 1.08; 95% CI: 1.01- 1.15; p = 0.01). Multiple sensitivity analyses further reinforced the validity of these findings. Multivariable Mendelian randomization adjusting for gestational hypertension confirmed the independent effect of gestational diabetes mellitus on postpartum depression and vice versa for gestational hypertension.

**Conclusion:** Both gestational diabetes mellitus and gestational hypertension increase the incidence of postpartum depression. By focusing on interventions to manage these prenatal conditions, nursing professionals can play a crucial role in potentially reducing the incidence of postpartum depression.

## Introduction

The diagnostic criteria for postpartum depression (PPD) are the same as for non-perinatal major depression, encompassing symptoms such as depressed mood, loss of interest, anhedonia, sleep and appetite disturbance, impaired concentration, psychomotor disturbance, fatigue, feelings of guilt or worthlessness, and suicidal thoughts, with prevalence rates ranging from 13-19% [1,2]. A major depressive episode “with peripartum onset” occurs “during pregnancy or in the four weeks following delivery”, as per the American Psychiatric Association’s Diagnostic and Statistical Manual of Mental Disorders [3]. In clinical practice and research, PPD is often more broadly defined as occurring from 4 weeks up to 12 months after childbirth [4]. PPD has significant negative consequences for both mothers and their children. These consequences include maternal suffering, impaired mother-infant relationships, and long-term negative impacts on child development across cognitive, behavioural, and social-emotional domains [5].

Gestational diabetes mellitus (GDM) and hypertensive disorders of pregnancy (HDP), including gestational hypertension (GH) and preeclampsia, are common pregnancy complications characterized by glucose intolerance and elevated blood pressure, respectively. Previous observational studies have shown that both GDM and GH are associated with several adverse maternal and fetal outcomes, including an increased risk of PPD. For instance, GDM, which affects 5.8-12.9% of the global population [6], increased the risk of PPD symptoms by 32% [7]. Similarly, GH significantly increased the risk of PPD, with an incidence rate ratio (IRR) of 1.84 (95% CI: 1.33-2.55) [8]. The causal pathways remain unclear despite these associations due to potential confounding factors in observational studies.

MR offers a robust methodological approach to overcome the limitations of observational studies and establish a causal relationship [9]. MR uses genetic variants as instrumental variables to infer causality, minimizing biases from confounding variables and reverse causation. This method leverages the random assortment of genes at conception to provide a more reliable estimate of the causal effects of GDM and GH on the risk of PPD [10].

In nursing practice, understanding the implications of GDM and GH on maternal mental health is crucial. By identifying these risk factors, nurses can provide targeted support and early interventions, improving outcomes for both mothers and infants. This essay examined the causal relationships between GDM, GH, and PPD through a MR approach, offering insights into nursing care and preventive strategies.

## Methods

### Study design

MR analysis investigated the causal relationships between risk factors and diseases using publicly available data from genome-wide association studies (GWAS). In this context, genetic variations were treated as instrumental variables, helping to reduce the impact of unmeasurable confounding factors and thus leading to more robust causal inferences. MR was based on the following three core assumptions: (1) genetic instrument variables (GIVs) were strongly associated with the exposure; (2) GIVs were independent of confounders, known or unknown; and (3) GIVs solely affected outcome risk through exposure, not other pathways [11]. **Fig 1**. provides an overview of the MR research framework. No additional ethical approval was required since the data was publicly available and relevant research had already undergone ethical review.

**Fig. 1.**
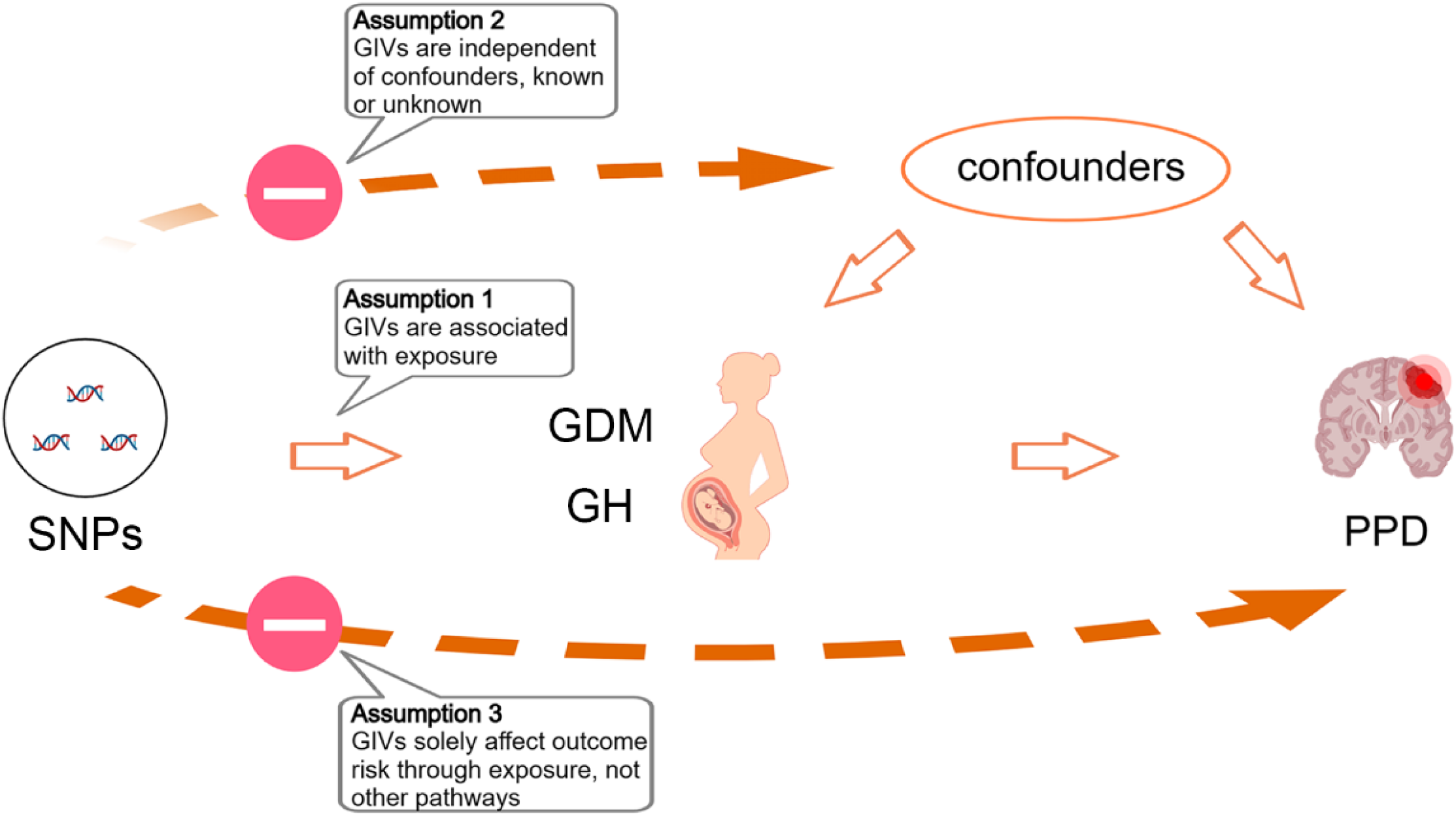
The Mendelian rando1nisation fran1ework shows a causal relationship between GDM, GH, and PPD. GDM, gestational diabetes mellitus; GH, gestational hypertension; PPD, postpartum depression; SNPs, single nucleotide polymorphisms; GIVs, genetic instrument variables

### Data sources

We retrieved data from the GWAS database (https://gwas.mrcieu.ac.uk/), which included GDM (123,579 individuals, finn-b-GEST_DIABETES), GH (123,579 individuals, finn-b-O15_HYPTENSPREG), and PPD (67,205 individuals, finn-b- O15_POSTPART_DEPR). The remaining relevant information could be found in **Table 1**. We used a rigorous quality control method to select GIVs, namely single nucleotide polymorphisms (SNPs). Based on MR analysis methods, a minimum of 10 SNPs was required [12]. Using a threshold of clump p = 1×10^-5^, we selected 34 SNPs associated with GDM and 45 SNPs associated with GH [13]. We set parameters kb = 10,000 and r2 = 0.001 to eliminate linkage disequilibrium between SNPs. We calculated the F-statistic to assess the risk of weak instrument bias using the formula F = [(n-k- 1)/k] × [R^2^/(1-R^2^)], where n is the sample size, k is the number of SNPs, and R^2^ represents the proportion of variance explained by each SNP [14]. If the F-statistics for all SNPs were greater than 10, it indicated a low risk of weak instrument bias [10].

**Table 1.** Information on data sets.

|  | Disease | Sample size | Number of SNPs | Population | Origin |
| --- | --- | --- | --- | --- | --- |
| Exposure | GDM | 123,579 | 16,379,784 | European | FinnGen |
|  | GH | 123,579 | 16,379,784 | European | FinnGen |
| Outcomes | PPD | 67,205 | 16,376,275 | European | FinnGen |

GDM, gestational diabetes mellitus; GH, gestational hypertension; PPD, postpartum depression; SNPs, single nucleotide polymorphisms.

### Mendelian randomization analyses

The causal relationship between exposure and outcome was evaluated using five methods: inverse variance weighted (IVW) [13], MR-Egger [15], weighted median [16], simple mode, and weighted mode [17]. IVW was the primary method, requiring all SNPs to adhere to the three MR principles. It does not consider an intercept in regression and uses the inverse of the outcome variance (square of the standard error) as a weight for fitting. MR-Egger also employs the inverse of the outcome variance as weights but includes an intercept, mainly aiming to assess the presence of horizontal pleiotropy. The weighted median sorts all SNP effects by magnitude and takes the median value as the final effect estimate. Simple mode treats the association of each instrumental variable with both the exposure and the outcome as having equal weight. It is applicable when there are only a few instrumental variables. Weighted mode weights each instrumental variable based on its association strength with the exposure, allowing for a more accurate estimation of causal effects. In the absence of heterogeneity and horizontal pleiotropy, IVW estimates are preferred. When only heterogeneity is present without horizontal pleiotropy, the IVW random effects model can be used. We used the MR-Steiger test to verify the direction of causality. p < 0.05 indicated the absence of reverse causality. The statistical power was calculated using prior observational studies with mRnd (https://shiny.cnsgenomics.com/mRnd/) [18].

### Analysis of heterogeneity and horizontal pleiotropy

Sensitivity analyses were conducted to assess the robustness of the research findings and potential biases. Heterogeneity may arise from genetic and environmental differences, measurement bias, interactions, and sample variations. The Cochran’s Q statistic was used to evaluate whether there was heterogeneity in the IVW and MR-Egger results, with p > 0.05 indicating no heterogeneity among SNPs [19]. The intercept term in the MR-Egger method was used to indicate horizontal pleiotropy. If there was a significant difference between this intercept term and 0 (p < 0.05), it suggested the presence of horizontal pleiotropy. Additionally, we employed MR-PRESSO to detect horizontal pleiotropy, where p > 0.05 implied that the impact of horizontal pleiotropy on the results could be disregarded [20]. A funnel plot was generated using the mr_funnel_plot function. If the SNPs are symmetrically distributed around the IVW in a funnel shape, it indicates the absence of significant horizontal pleiotropy. Lastly, we used the leave-one-out approach to iteratively remove one SNP at a time and calculated the meta-effect of the remaining SNPs. If the overall error line does not change significantly and all errors fall on the same side of 0 after removing each SNP, the results are stable and not influenced by any SNP [21]. All the analyses above were performed using the TwoSample MR package (version 0.5.7) in R (version 4.3.1).

## Results

### Gestational diabetes mellitus and postpartum depression

In the analysis of PPD, all 34 SNPs correlated with GDM were examined **(S1 Table)**. These SNPs displayed a robust F statistic exceeding 10 (mean = 54.67, range = 45.22 - 184.10), suggesting a minimal likelihood of weak instrument bias.

The IVW indicated a significant positive association, with an OR of 1.09 (95% CI: 1.03 - 1.14; p = 1.24×10^-3^). Weighted median yielded comparable results, demonstrating an OR of 1.11 (95% CI: 1.02 - 1.21; p = 0.01). Weighted mode indicated a significant positive association, with an OR of 1. 11(95% CI: 1.01 - 1.23; p = 0.03) **(Fig 2a)**. The beta values from MR-Egger and simple mode were consistent with the direction of those from IVW, weighted median, and weighted mode **(Figs 3a and 4a)**. The Cochran’s Q test did not find any significant evidence of heterogeneity between the SNPs (p = 0.58). The MR-Egger intercept test (p = 0.57) and MR-PRESSO analysis (p = 0.57) indicated no evidence of horizontal pleiotropy. SNPs were symmetrically distributed around the IVW in a funnel shape, indicating no significant horizontal pleiotropy **(Fig 5a)**. The leave-one-out analysis showed that the causal relationship remained stable when any single SNP was removed **(Fig 6a)**. The MR-Steiger test showed that the influence of GDM on PPD risk was the correct causal direction (p = 5.07×10^-39^). Our study had a power of 100% for detecting the causal effect of GDM on PPD.

**Fig. 2.**
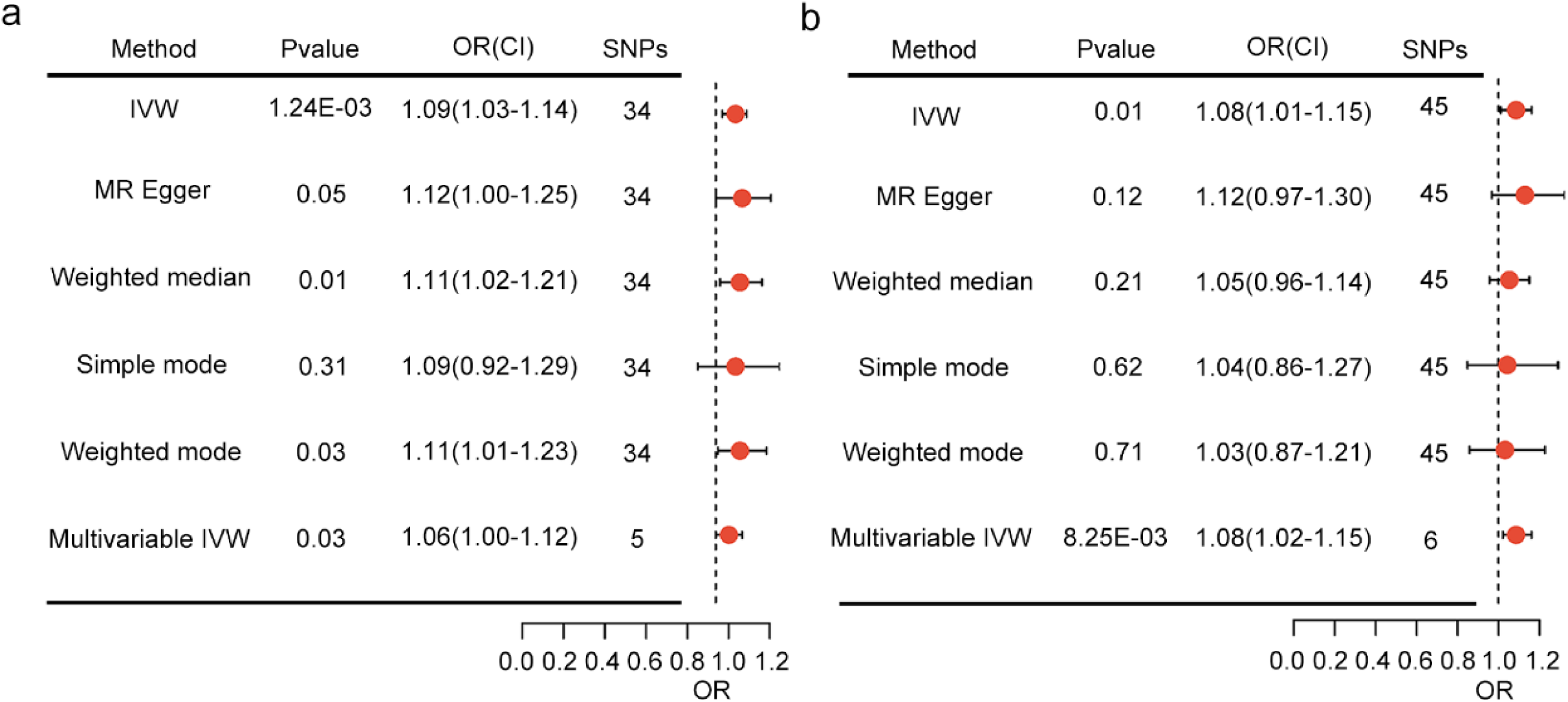
The OR and CI presented in the study demonstrate the impact of GDM and GH on the risk of PPD. The study compares the outcomes of MR analyses carried out using odds ratios; CI, confidence intervals

**Fig. 3.**
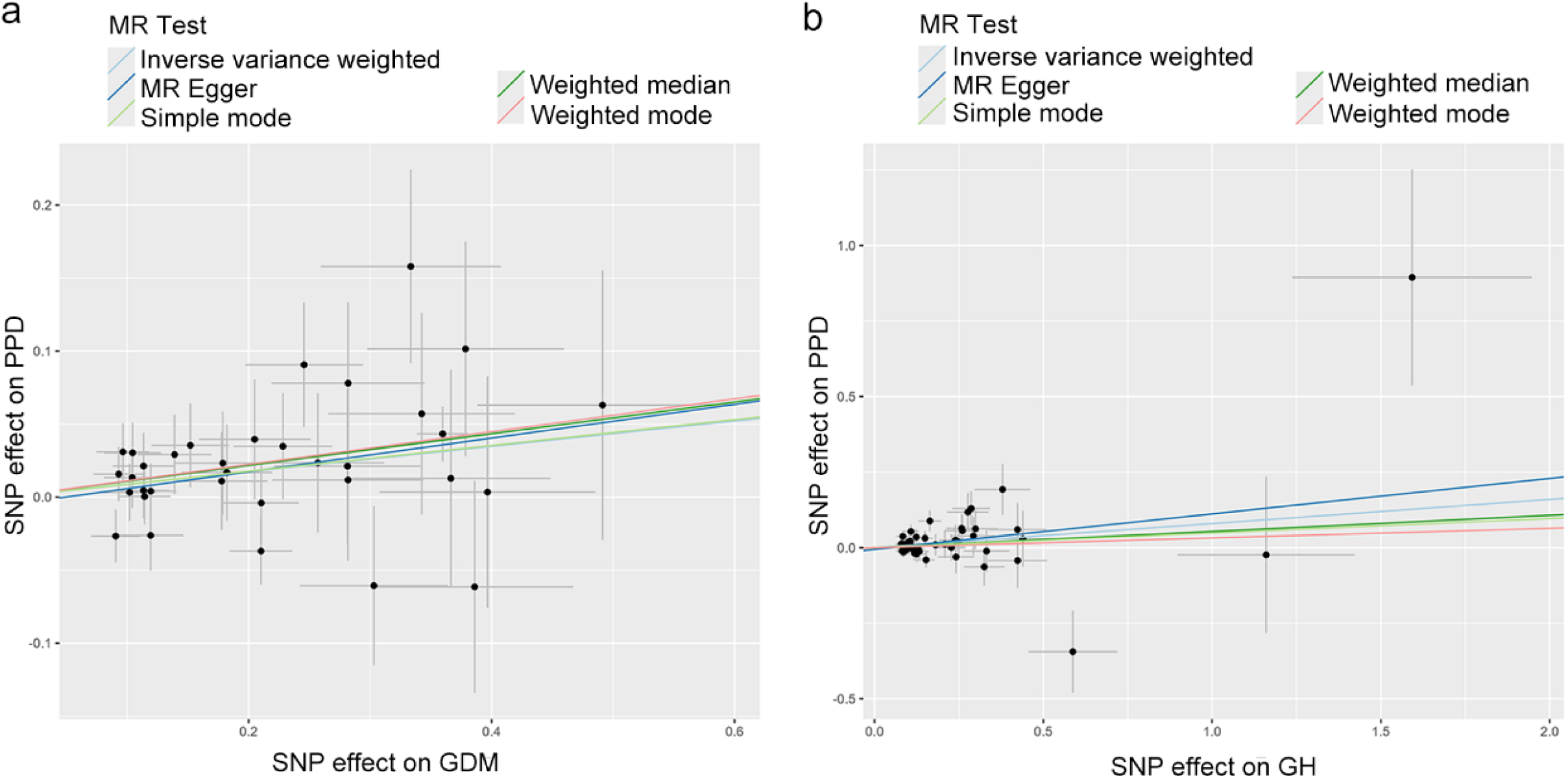
The scatter plot showcases the effect of GDM and GH associated with SNP on PPD, represented on the log-odds scale. The lines’ slopes indicate the causal association for each method

**Fig. 4.**
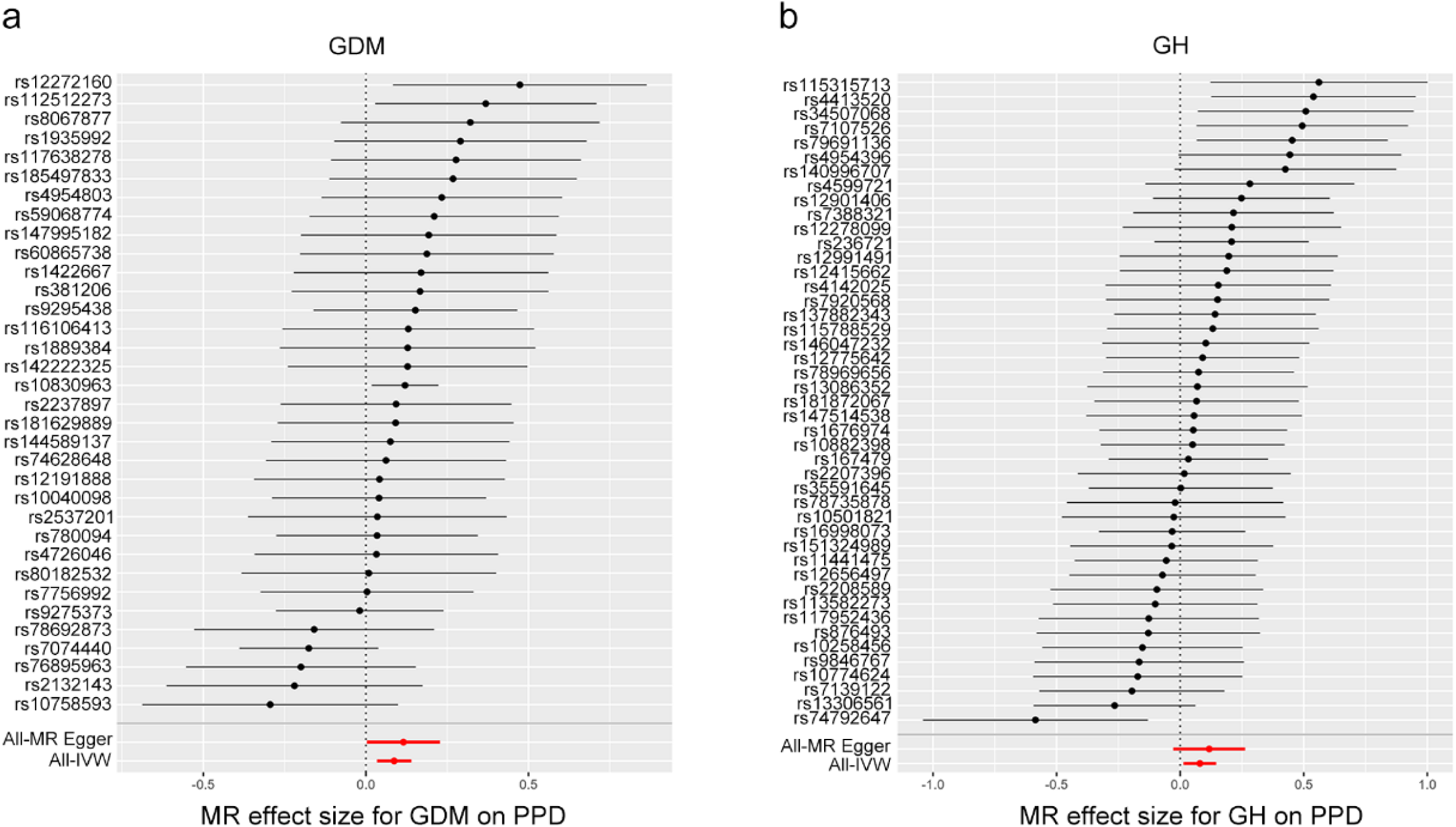
A forest plot is utilized tovisually represent thecausal effect of each SNP on the risk of PPD

**Fig. 5.**
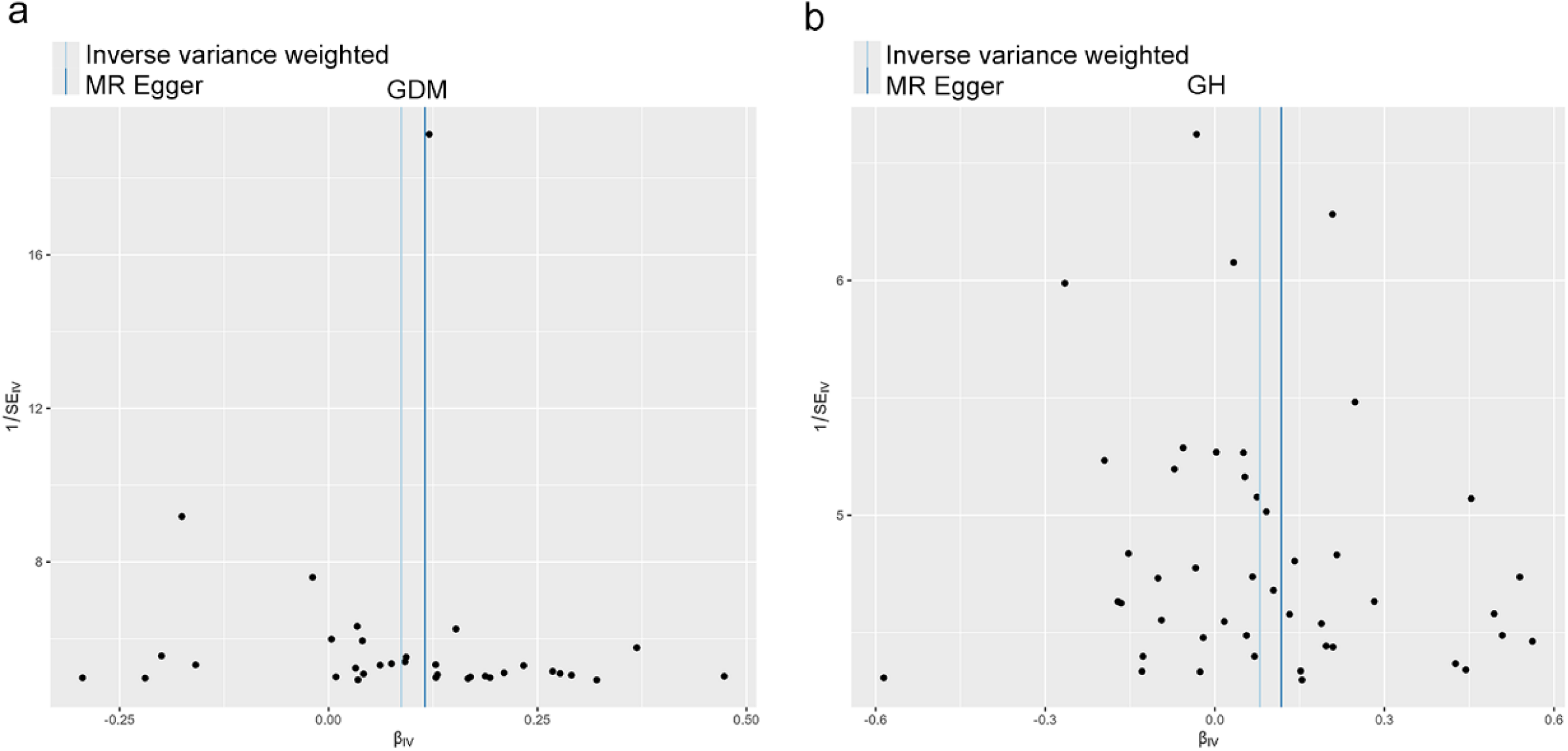
Funnel plots are utilized to visually represent the general heterogeneity of MR estimates regarding the impact of GDM and GH on the risk of PPD

**Fig. 6.**
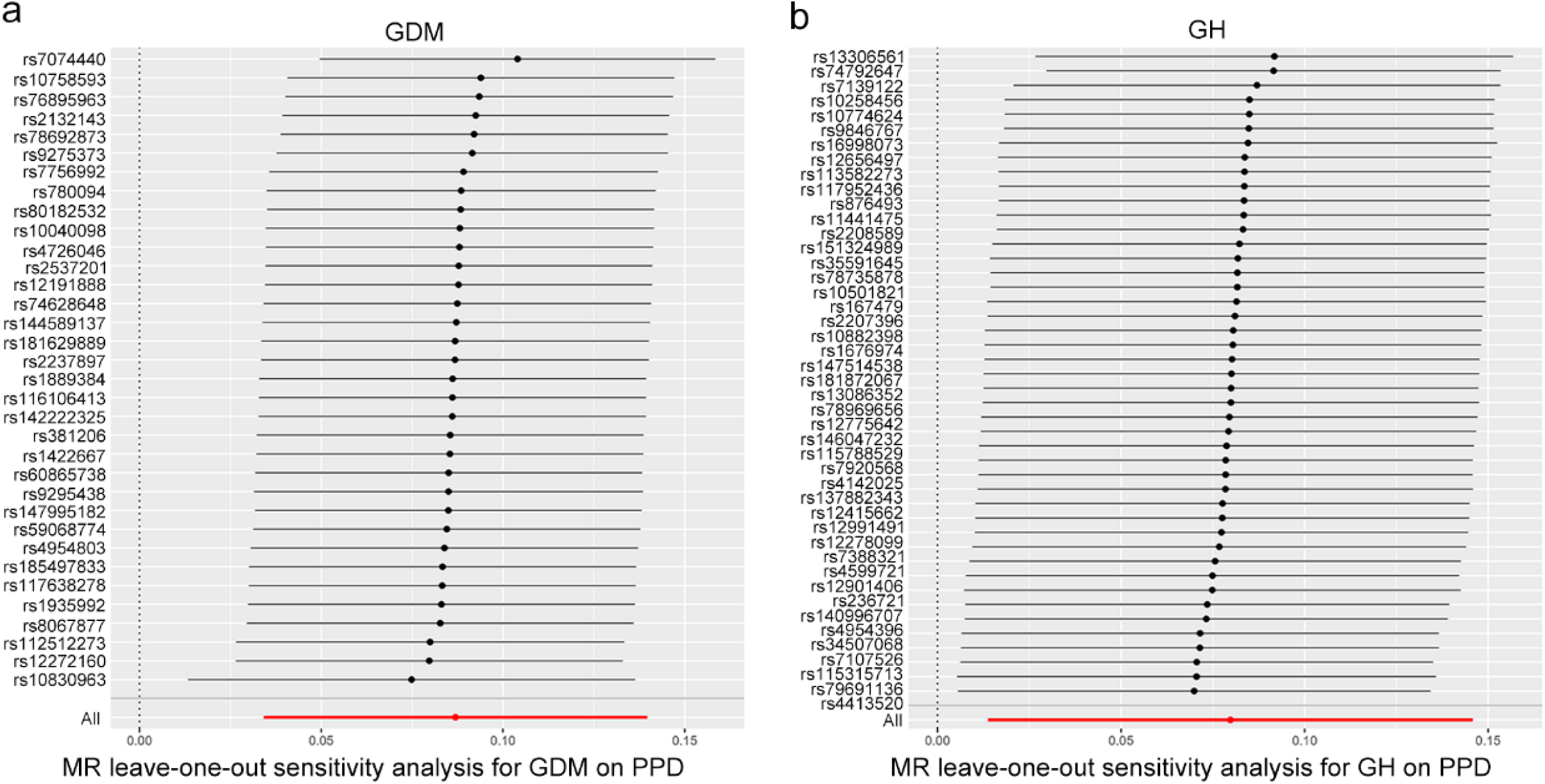
A leave-one-out plot is generated to visually illustrate the causal effect of GDM and GH on the risk of PPD by systematically excluding one single nucleotide polymorphism at a time

In the multivariable MR analysis that adjusted for preeclampsia, significant evidence indicated a direct causal relationship between GDM and an increased risk of PPD (IVW OR = 1.06, 95% CI: 1.00 - 1.12, p = 0.03) **(Fig 2a)**.

### Gestational hypertension and postpartum depression

The summary statistics for PPD included all 45 independent SNPs associated with GH **(S2 Table)**. The F statistic of these SNPs was > 10 (mean = 38.93, range = 34.26 - 54.39), indicating a low risk of weak instrument bias.

This study demonstrated a positive association between genetically predicted GH and PPD, with an OR of 1.08 (95% CI: 1.01 - 1.15, p = 0.01) in the IVW analysis **(Fig 2b)**. The beta values of MR-Egger, weighted median, simple mode, and weighted mode were consistent with the direction of the beta value of IVW **(Figs 3b and 4b)**. No statistical evidence of heterogeneity was found across instruments (p = 0.15). The MR-Egger intercept test (p = 0.57) and MR-PRESSO analysis (p = 0.18) showed no horizontal pleiotropy. The symmetric funnel-shaped distribution of SNPs around the IVW indicated no significant horizontal pleiotropy **(Fig 5b)**. In addition, no single SNP affected the overall estimate, as shown by the leave-one-out analysis **(Fig 6b)**. The MR-Steiger test confirmed that GH influences the risk of PPD in correct causal direction (p = 2.88×10^-45^). Our study had 100% power to detect the causal effect of GH on the risk of PPD.

In the multivariable MR analysis, after adjusting for GDM as a potential confounder, the causal effect of GH on the risk of PPD was still observed (IVW OR = 1.08, 95% CI: 1.02 - 1.15, p = 8.25×10^-3^) **(Fig 2b)**.

## Discussion

This study employed the MR method to estimate the causal effects of GDM and GH on the risk of PPD. By utilizing both univariable MR and multivariable MR, we identified an independent causal impact of genetically predicted GDM on the risk of PPD (IVW OR = 1.09, 95% CI: 1.03 - 1.14, p = 1.24×10^-3^), and found clear evidence of an independent causal effect of GH on PPD risk (IVW OR = 1.08, 95% CI: 1.01 - 1.15, p = 0.01).

Previous observational studies have shown that both GDM and GH increase the risk of PPD. In a 4-year cohort study involving 2,477 females, GDM was linked to a 4.62-fold increased risk of PPD (95%CI 1.26 - 16.98) [22]. Similarly, a study from the Kuopio Birth Cohort, encompassing 1,066 women without prior mental health concerns, found GDM increased the likelihood of PPD symptoms with an OR of 2.23 (95%CI 1.23 - 4.05) after adjusting for various maternal and neonatal factors [23]. This suggested that GDM was a potential independent risk factor for PPD. A separate cross-sectional study in Brazil between 2015 and 2017, with 168 postpartum women, found that 23.8% displayed depressive symptoms, with a significant correlation between PPD and HDP (Spearman [rs] 0.219; P = 0.004) [24]. Lastly, in the Rhea mother-child cohort in Greece, comprising 1,037 women, GH was significantly linked to increased PPD symptoms (β coefficient 1.86, 95% CI 0.32 - 3.41) [25]. However, previous observational studies had limitations, such as small sample sizes and confounding factors, necessitating a more robust method to infer the causal relationship between GDM, GH, and PPD risk.

Our MR study offered a novel and comprehensive evaluation of the causal link between GDM, GH, and PPD. Unlike traditional observational epidemiological approaches, MR can more robustly suggest causality, potentially circumventing the pitfalls of unmeasured confounders and reverse causation. The causative estimates derived from our analysis aligned with previous observational studies on GDM, GH, and PPD. Utilizing two GWAS datasets, we pinpointed 34 and 45 SNPs and employed five models: IVW, weighted median, MR-Egger, simple mode, and weighted mode. Our IVW analysis revealed that GDM and GH were risk factors for PPD, with a 1 SD increase in GDM leading to a 9% increased risk of PPD, as determined by weighted median and weighted mode. Additionally, we found that for every 1 SD increase in GH, the risk of PPD increased by approximately 8%.

The mechanisms underlying the relationship between GDM, GH, and the risk of PPD remain incompletely understood. GDM may act as a physiological stressor, triggering neuroendocrine dysregulation and inflammatory responses that predispose women to depressive symptoms [2]. Insulin resistance, a hallmark of GDM, has been associated with altered neurotransmitter function, potentially contributing to mood disturbances [26]. Metabolic perturbations accompanying GDM could disrupt the hypothalamic-pituitary-adrenal (HPA) axis, leading to aberrant cortisol signaling implicated in depression pathophysiology [27]. A recent rat study demonstrated that GDM disrupts tryptophan metabolism and alters gut microbiota composition, providing a putative physiological basis for PPD [28]. Women who experience pregnancy complications, such as hypertension, are at an increased risk of physical morbidity in the postpartum period, which may contribute to elevated rates of PPD [25]. Preeclampsia is characterized by excessive inflammatory responses, with elevated levels of pro-inflammatory cytokines like IL-6, IL-8, TNF-alpha, and C-reactive protein associated with PPD development [29–33]. Genetic factors may also play a role, as specific polymorphisms, such as MTHFR C677T, 5-HTT, and ESR, are linked to increased susceptibility to preeclampsia and PPD [34–37]. Alterations in folate metabolism due to MTHFR gene variants may influence serum folate levels, potentially increasing the risk of PPD [38,39].

A key strength of our study was utilizing a two-sample Mendelian randomization design, which mitigates potential biases from confounding when assessing the causal relationships between GDM, GH, and PPD. Additionally, our approach may have minimized the risk of population stratification bias by focusing solely on individuals of European ancestry.

Our study had several limitations. Primarily, participants were exclusively sourced from a European ancestry GWAS database, necessitating validation of causal associations in more diverse populations. Additionally, the absence of weight-stratified summary data for GDM and GH exposures hinders the examination of their relationship with PPD risk. Factors such as age and pre-existing health conditions could confound our results; however, the limited genetic data on these variables posed challenges for comprehensive adjustments.

Future research should prioritize understanding the biological mechanisms underlying the causal relationships between GDM, GH, and PPD. Investigations into neuroendocrine dysregulation and inflammatory responses could provide insights into these pathways. Longitudinal cohort studies are needed to track the progression from GDM and GH to PPD over time, which would clarify temporal dynamics and mediating factors. Expanding research to diverse populations will help ensure the findings’ generalizability. Additionally, randomized controlled trials should test targeted interventions for managing GDM and GH during pregnancy to assess their effectiveness in reducing PPD incidence. Further genetic studies with larger GWAS datasets could identify additional SNPs, enhancing our understanding of genetic risk factors.

Updating prenatal care guidelines to include routine screening for GDM and GH is essential for comprehensive assessments that could reduce postpartum depression PPD risk. Integrated care models, involving obstetricians, endocrinologists, and mental health professionals, should be developed to offer holistic care and timely interventions.

Enhancing education and training for healthcare professionals, particularly nurses and midwives, is crucial to recognizing and addressing GDM, GH, and PPD early. Public health campaigns can raise awareness of the risks associated with these conditions, encouraging early care and adherence to management plans. Additionally, policy development should focus on funding research in maternal health, emphasizing preventing and managing GDM, GH, and PPD through innovative projects and improved healthcare infrastructure.

## Conclusions

Our two-sample MR analysis offered genetic evidence supporting the causal role of GDM and GH in the onset of PPD. These findings underscored the necessity for nurses and healthcare professionals to provide tailored clinical care, emphasizing education, support, and early intervention to mitigate PPD risk in individuals with a history of GDM and GH.

## Data Availability

All relevant data are within the manuscript and its Supporting Information files.

https://gwas.mrcieu.ac.uk/

## Acknowledgements

We thank Associate Professor Rongzu Nie from Zhengzhou University of Light Industry for his valuable suggestions on the manuscript.

## Supporting information

S1 Table. Independent SNPs associated with GDM

S2 Table. Independent SNPs associated with GH

